# Immunosurveillance of CCR6+ T-cells predicts treatment response to dimethyl-fumarate: implications for personalized treatment strategies in multiple sclerosis

**DOI:** 10.1101/2020.05.15.20102137

**Authors:** Constantinos Alifieris, Seunghee Kim Schultze, Ilana Katz Sand, Patrizia Casaccia, Achilles Ntranos

## Abstract

**Objective:** The field of multiple sclerosis (MS) has seen a tremendous expansion of treatments in the past decade. However, treatment response in individual patients can currently be determined only by waiting for breakthrough disease activity to occur. This highlights a critical need for biomarkers that can predict treatment response and stratify the risk of impeding disease activity before damage is inflicted to the CNS. Here we show that CCR6+CD3+ T-cell surveillance in peripheral blood can be used to discriminate responders and non-responders to dimethyl-fumarate.

**Methods:** A cohort of 101 treatment-naïve, dimethyl-fumarate (DMF) treated MS patients and healthy controls was immunophenotyped and then responders and non-responders were determined retrospectively after clinical and radiographic follow up. Receiver operating characteristic (ROC) curve, linear and logistic regression, mixed effects models, and cox proportional hazards were used for the analysis.

**Results:** Among various clinical and immunophenotypic metrics, the percentage of CCR6+CD3+ T-cells was the most significant predictor of impending disease activity. This immunophenotypic metric was able to discriminate responders and non-responders to DMF with an area under the ROC of 0.85 (95% CI: 0.71–0.99), which was higher than that achieved using surrogate metrics for T-helper-1-like T-helper-17 or T-cytotoxic-17 cells. DMF-treated patients with the highest percentage of CCR6+CD3+ T-cells had a significantly higher risk of impending disease activity compared to patients with a low percentage.

**Interpretation:** Changes in CCR6+CD3+ T-cells in the periphery could precede disease activity by many months and potentially serve as an early biomarker of treatment response, at least for DMF. These results have implications for novel personalized treatment strategies in MS.

## Introduction

The field of multiple sclerosis (MS) has seen a tremendous expansion of available treatments in the past decade, but progress regarding biomarkers to predict treatment success has lagged. In current practice, measurement of treatment efficacy involves waiting for breakthrough disease activity to occur, which may lead to accumulation of permanent disability. There is therefore a significant clinical need to develop biomarkers of treatment response in MS patients that can stratify the risk of treatment failure and impending breakthrough disease activity before it occurs.

We know that relapses in MS can be prevented by blocking the trafficking of peripheral immune cells to the central nervous system, suggesting that changes in the periphery could potentially precede disease activity. We hypothesized that the presence or absence of a peripheral immunologic response to a drug could be used to predict clinical response to it in MS patients. Based on our previous work, dimethylfumarate (DMF), an effective immunomodulatory therapy for MS, can reduce the percentage of CCR6+CD4+ and CCR6+CD8+ T-cells by epigenetically modulating microRNA-21 expression^1^. CCR6 is known to be highly expressed on T-helper-17 (Th17) and T-helper-1-like Th17 cells (Th1.17), as well as IL17+CD8+ cytotoxic T-cells (Tc17). CCR6 expression allows these cells to enter the uninflamed CNS and initiate disease in mouse models of MS^2–5^, which suggests that CCR6 is a brain-homing chemokine receptor. CCR6 has also been found to be expressed in myelin-reactive T-cells from the blood of MS patients, as well as CD4+ and CD8+ T-cells in the CSF of MS patients^6–8^. This prompted us to test whether this mechanistic immunological metric could predict future breakthrough disease activity in patients with MS treated with DMF. To this end, we utilized a cohort of a total of 101 treatment-naïve and DMF-treated MS patients and healthy controls, who underwent immunophenotyping and then retrospectively classified as responders and non-responders based on real world clinical and radiographic follow-up to determine the development of disease activity. Our results show that surveilling the peripheral CCR6+ T-cell percentage provides a mechanistic immunophenotypic metric that can predict clinical response of DMF-treated patients and separate responders from non-responders before the CCR6+ T-cells as a biomarker of DMF response in MS development of disease activity. These results suggest that peripheral immune surveillance could be potentially used as a personalized decision-making strategy in MS patients.

## Methods

### Study design and Participants

This research was approved by Mount Sinai’s Institutional Review Board (IRB) and written informed consent was obtained for all patients according to the Declaration of Helsinki. Inclusion criteria were age between 18 and 65 years-old, diagnosis of Relapsing-Remitting Multiple Sclerosis (RRMS) by McDonald 2010 criteria^9^ and being treatment-naive or on DMF treatment for at least three months. DMF 240 mg twice daily was given orally. Exclusion criteria included high dose steroids within one-month, immunosuppressant medications or antibiotics within three months and a diagnosis of diabetes or inflammatory bowel disease. The cohort was assembled by consecutive sampling between 2014 and 2016, and was clinically followed for 2 years after the last blood draw. A subgroup of DMF treated patients also had their blood drawn before starting DMF therapy, as treatment naïve. Age, self-reported gender and race, body mass index (BMI), disease duration, Expanded Disability Status Scale (EDSS) and treatment duration were collected at baseline. Treatment duration was defined as the time when patients were first given DMF to the day of the index test. Responders and non-responders were identified retrospectively based on no evidence of disease activity (NEDA-3)^10,11^ by reviewing clinical and radiographic data from up to 24 months of follow-up after their blood draw at a comprehensive MS center (Table 1).

**Table 1.** Patient demographic and disease characteristics.

|  | Healthy controls<br>(HC)<br>N=10 | Naïve<br>(Na)<br>N=43 | Subjects RRMS<br>Cross-sectional |  | Stats | Subjects RRMS<br>Longitudinal<br>Sub-analysis |  | Stats |
| --- | --- | --- | --- | --- | --- | --- | --- | --- |
|  |  |  | Responders<br>(R)<br>N=39 | Non-Responders<br>(NR)<br>N=9 |  | Responders (R)<br>N=10 | Non-Responders<br>(NR)<br>N=3 |  |
| Age<br>(SD) | 30.6<br>(7.8) | 36.63<br>(10.54) | 43.32<br>(8.05) | 37.54<br>(13.12) | ANOVA<br>p=0.0001<br>Tukey:<br>RvsNa,<br>RvsHC<br>p<0.0001 | 43.09<br>(8.31) | 38.28<br>(7.87) | Welch t-test<br>p=0.4247 |
| Gender | F: 9 (90%) | F: 29 (67%) | F: 24 (62%) | F: 4 (44%) | chi-squared<br>p=0.2029 | F: 6 (60%) | F: 1 (33%) | chi-squared<br>p=0.495 |
| Race | Caucasian: 7<br>(70%)<br>NonCaucasian: 3 | Caucasian: 29<br>(67%)<br>NonCaucasian: 14 | Caucasian: 29<br>(74%)<br>NonCaucasian: 10 | Caucasian: 5<br>(56%)<br>NonCaucasian: 4 | chi-squared<br>p=0.7195 | Caucasian: 5<br>(50%)<br>NonCaucasian: 5 | Caucasian: 1<br>(33%)<br>NonCaucasian: 2 | chi-squared<br>p=0.8015 |
| BMI<br>(SD) | NA | 25.48<br>(5.67) | 25.67<br>(5.01) | 26.92<br>(5.05) | ANOVA<br>p=0.762 | 27.63<br>(5.31) | 27.21<br>(4.99) | Welch t-test<br>p=0.9076 |
| Disease<br>Duration-<br>months<br>(SD) | 0 | 37.88<br>(51.86) | 111.07<br>(92.93) | 82.42<br>(51.13) | ANOVA<br>p=0.001<br>Tukey:<br>RvsNa<br>p<0.001 | 39.99<br>(15.97) | 41.54<br>(3.90) | Welch t-test<br>p=0.7838 |
| EDSS<br>Median<br>(Range) | 0 | 1.5<br>(0-3.5) | 1.14<br>(1.04) | 1.61<br>(1.17) | ANOVA<br>p=0.325 | 1.5<br>(0-3) | 1<br>(1-2) | Welch t-test<br>p=0.5162 |
| Treatment<br>Duration-<br>months<br>(SD) | 0 | 0 | 14.98<br>(8.07) | 17.78<br>(10.41) | ANOVA<br>p=0.379 | 9.6<br>(4.4) | 6.3<br>(5.7) | Welch t-test<br>p=0.4349 |

### Immunophenotyping

Peripheral blood was collected and processed fresh within three hours by the Human Immune Monitoring Core facility at Mount Sinai. Cells were stained with a pre-optimized cocktail of antibodies against CD45, CD3, CD4, CD8, CCR4 and CCR6 as previously described^1^. Of note, since the flow cytometry panel design occurred before the first reports of memory T-cell reduction by DMF, CD45RO and CCR7 were not included in the cocktail of antibodies. Cells were then acquired on a BD LSR Fortessa flow cytometer (BD, San Jose, CA). Post acquisition analysis of cell populations was done with FlowJo (Treestar Inc, San Carlos, CA) (Supplementary Figure 1).

### Test methods

The index test was the percentage of CCR6+CD3+ T-cells in the blood of MS patients. Positivity cutoff was determined based on Youden’s J statistic^12^. The reference standard was the development of clinical or radiographic disease activity based on NEDA-3. Relapse information was abstracted from the medical record retrospectively. Each chart was reviewed to ensure that the relapse description met the usual definition of new or worsening neurological deficit lasting at least 24 hours in the absence of fever or infection. All clinical relapses were confirmed with imaging. The opinion of the treating MS specialist, who was blinded to the index test results, was not altered retrospectively. Radiographic relapse was defined as a new or clearly enlarging T2 lesion or T1 enhancing lesion as reported in the radiology report and/or MS specialist note and was confirmed on imaging. The neurological exam between visits was used to assess stability of EDSS. Relapse-free intervals were determined based on the time from last blood draw to clinical relapse or last known stable MRI (if there was a new non-enhancing T2 lesion) or one month prior to the detection of a new T1 enhancing lesion. We used the last observation carried forward method to impute missing data and six patients that did not complete 24 months of follow up, but had not exhibited disease activity for the time they were followed, were considered as responders. Sensitivity analysis revealed that the inclusion of all cases or only complete cases did not alter the results of this study. In fact, the exclusion of patients lost to follow-up results in similar or improved estimations of several metrics, including p-value of logistic regression, area under the receiver operating characteristic (ROC) curve (AUC) and cox-proportional hazard ratios. Therefore, to avoid any potential selection or attrition biases in our estimations, we included all patients in the following analyses.

### Statistical analyses

All statistical analyses were performed in R. The primary endpoint of this study was to assess the area under the ROC curve of CCR6+CD3+ T-cells between responders and non-responders to DMF. Given the retrospective nature of the study, a post-hoc power analysis would be uninformative^13,14^, but the ‘detectable’ AUC effect size could be determined when setting the power at 80% and the significance level at 5% and using the sample size of our cohort. The ‘detectable’ AUC effect size in this study given the above criteria is 0.79^15^ with a null hypothesis level of 0.5. Diagnostic accuracy was assessed by the area under the ROC curve and 95% confidence intervals were obtained with pROC^16^ in R. P-values from logistic regression analysis were obtained with the likelihood ratio test against a null model. Multiple hypothesis testing correction was performed, where appropriate, by controlling the false discovery rate^17^. Analysis of variance (ANOVA), Kaplan-Meyer and cox-proportional hazards analysis was performed in R^18^. An exploratory analysis of variability in diagnostic accuracy was done by separating the patients into two groups based on their baseline demographic characteristics (Supplementary Figure 2) and using a Bayesian bivariate hierarchical model (meta4diag) in R.

## Results

### Patient characteristics

We utilized immunophenotypic data from a cross-sectional cohort of 43 treatment-naïve, 48 DMF-treated relapsing remitting MS patients and 10 healthy donors (HD). 13 DMF treated patients had immunophenotyping before starting DMF treatment. The immunophenotyping analysis included the percentage of total CD3, CD4 and CD8 T-cells, as well as the percentage of those cells expressing CCR6^2,4^ and CCR4^19^. These subpopulations could be used as surrogates for Th17 (CD4+CCR4+CCR6+), Th1.17 (CD4+CCR4-CCR6+), Th2 (CD4+CCR4+CCR6-) and Tc17 (CD8+CCR6+) cells. We also analyzed CCR6+ CD3+CD4-CD8-double negative (DN) T-cells, total CD3-lymphocytes and CCR6+ CD3-CD4-CD8-lymphocytes (as a surrogate for CCR6+ B cells, since monocytes were excluded based on side scatter characteristics).

From 48 DMF-treated patients, two were completely lost to follow-up and four patients were lost to follow-up prior to 24 months. The average follow-up time of patients who did not complete 24 months was 5.49 (2.1–17) months (Figure 1). Treatment response (39 responders and 9 non-responders) based on NEDA-3 was determined retrospectively in a real-world setting, which included clinical and annual radiographic information within 24 months of follow up. From the subgroup of DMF patients who also had baseline immunophenotypic metrics, ten were responders and three were non-responders. The frequency of relapses in our cohort is in accordance with the relapse frequency of the two major clinical trials for DMF, DEFINE and CONFIRM (27% and 29% respectively)^20,21^. Patient demographic and disease characteristics were similar except for age and disease duration between responders and treatment-naïve patients of the cross-sectional cohort and healthy controls (Table 1).

**Figure 1.**
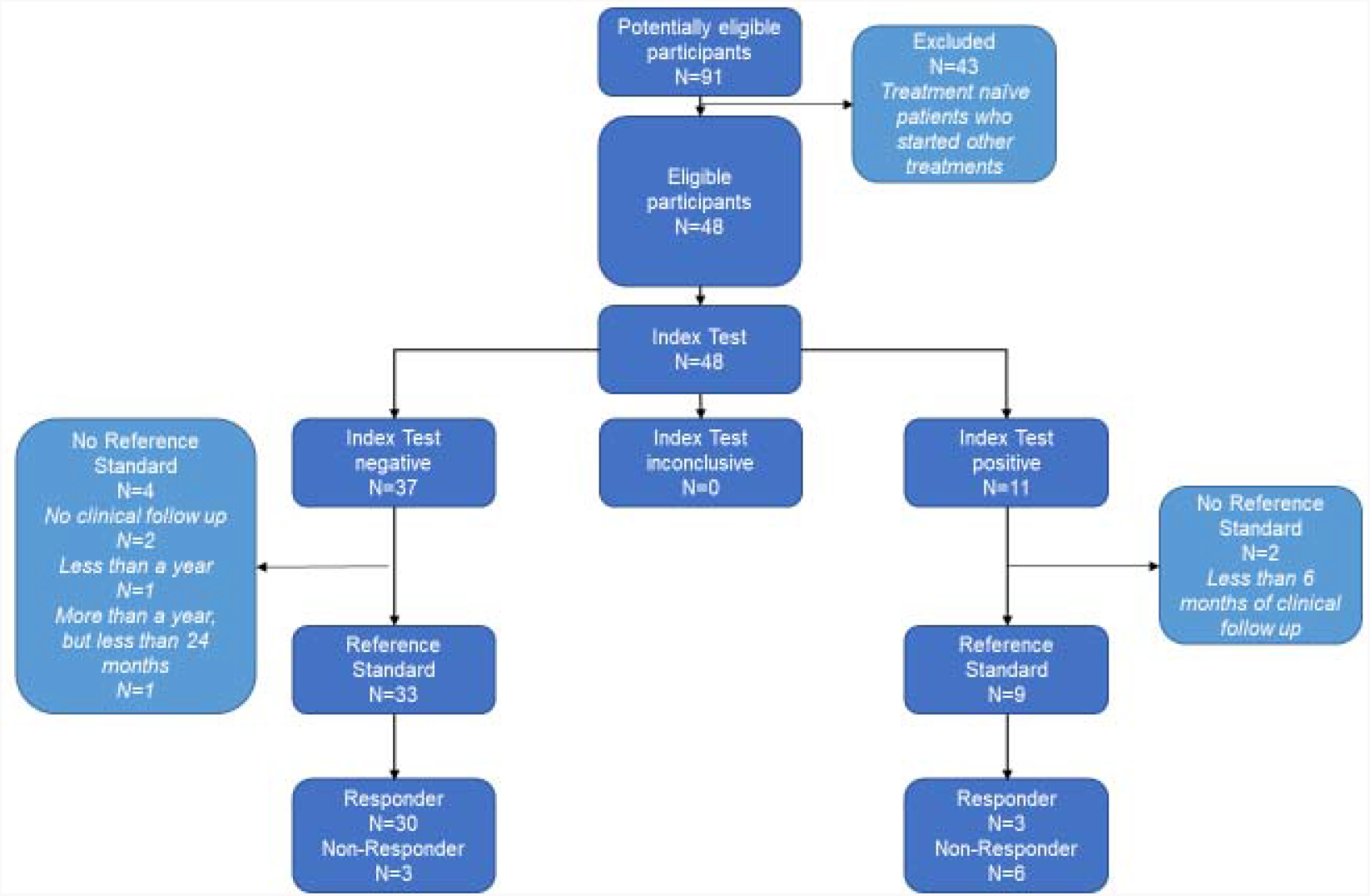
Study Flow Chart

### Responders and non-responders to DMF therapy differ in CCR6+ T-cell percentage

Our immunophenotyping analysis showed that responders and non-responders to DMF therapy differed the most on the percentage of CCR6+CD3+ T-cells, compared to the other T-cell subtypes that we measured (linear interaction model controlling for age, sex, race, baseline EDSS, disease duration and treatment duration, adj p<0.0001) (Figure 2A-B). Interestingly, the CCR6 percentage of DN T-cells (CD3+CD4-CD8-CCR6+) was associated with disease activity, but the percentage of CD3-CD4-CD8-CCR6+ lymphocytes, which should include CCR6+ B cells, was not (Figure 2A). Furthermore, CCR6+CD3+ T-cells were similar in responders and healthy donors (Figure 2C), suggesting a normalizing effect of the treatment. Non-responders, on the other hand, had similar percentage of CCR6+CD3+ T-cells with treatment-naïve patients, suggesting that either these non-responding patients did not experience a drop in their CCR6+CD3+ T-cells by DMF or they started with a higher percentage of these cells (Figure 2B-C). Indeed, based on our subgroup analysis of the 13 DMF treated patients who had immunophenotypic metrics before and after treatment, the reduction of CCR6+CD3+ T-cells was significantly lower in non-responders than in patients who had a good clinical response (linear mixed effect model, p=0.0235) (Figure 3A). The baseline percentage of CCR6+CD3+ T-cells appeared slightly higher in the non-responder group compared to responders, but this observation did not meet statistical significance with our current sample size (p=0.0697) (Figure 3B). Finally, after obtaining the percent change in CCR6+CD3+ T-cells between responders and non-responders, patients with future disease activity exhibited a lower percent change compared to stable MS patients (p=0.02472) (Figure 3C).

**Figure 2.**
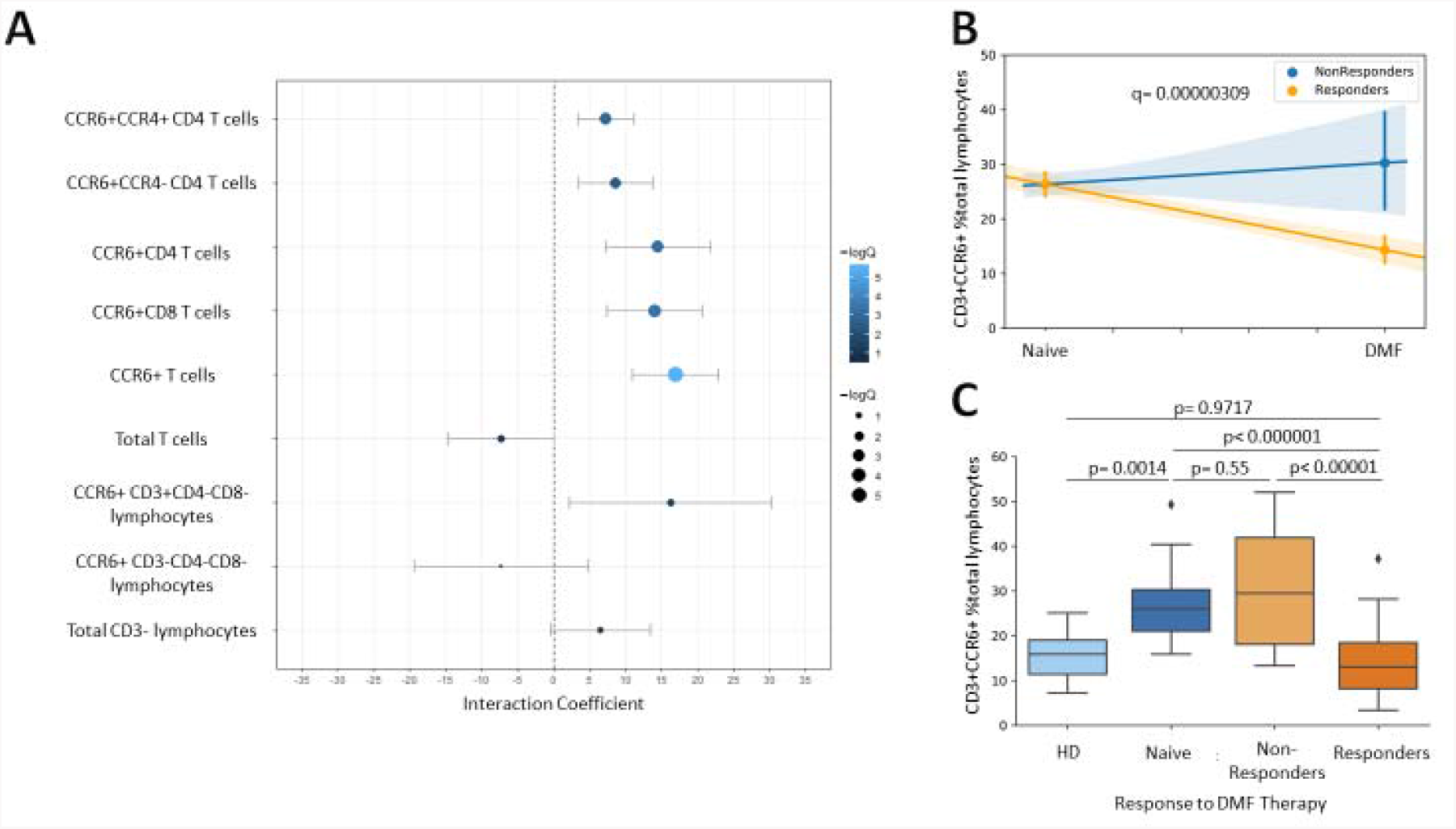
CCR6+ T-cells differ the most between responders and non-responders to DMF therapy: Immunophenotyping of peripheral blood was done in 43 treatment-naïve, 48 DMF-treated patients and 10 healthy donors (HD). Clinical follow up of patients treated with DMF revealed 39 responders and 9 non-responders to the drug. (A) Multivariate linear regression was used to analyze the relationship between various immunophenotypic metrics and the interaction between treatment status and the development of disease activity in these patients. The interaction coefficient from each of the models tested is depicted, with the size and color varying according to the negative logarithm of the adjusted p value (-logQ). The percentage of CCD6+CD3+ T-cells exhibited the strongest association with the development of future disease activity in DMF treated patients. (B) The CCR6+CD3+ T-cell interaction model between treatment status and future disease activity is shown. (C) The percentage of CCR6+CD3+ T-cells is shown among the different patient groups. The lines in the boxplots represent the quartiles of the dataset and the whiskers show the rest of the distribution except for outliers. Non-responders to DMF therapy have similar percentage with treatment-naïve patients, whereas responders to the drug have significantly lower CCR6+CD3+ T-cell percentage, which is similar to healthy donors. DN: double negative (CD3+CD4-CD8-)

**Figure 3.**
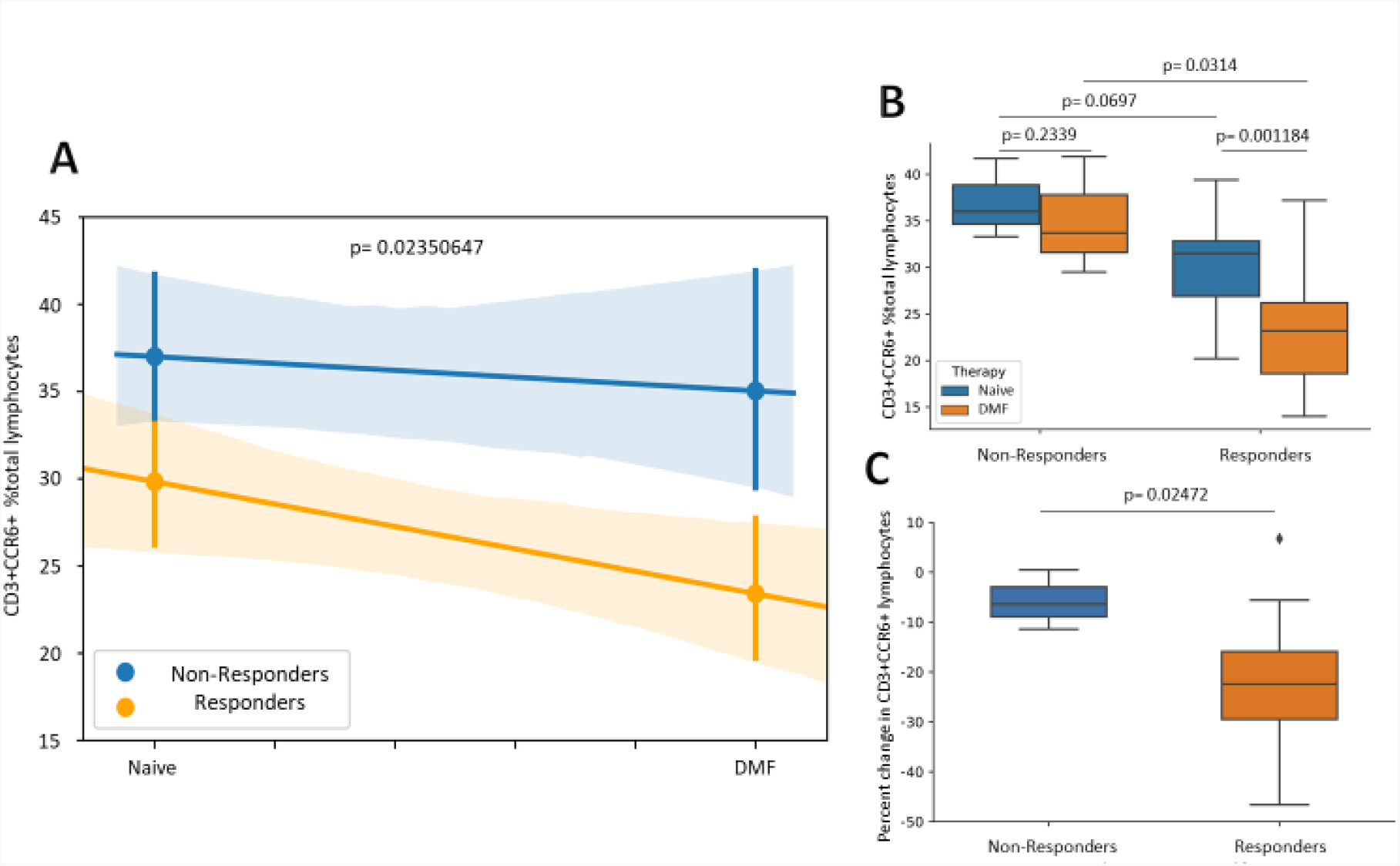
CCR6+ T-cell reduction after initiation of DMF is associated with good treatment response: Immunophenotyping was performed in 13 MS patients before and after treatment with DMF (A) The reduction dynamics of CCR6+CD3+ T-cell percentage were significantly different between responders and non-responders to DMF, with responders exhibiting stronger reduction in these cells compared to non-responders. (B) The percentage of CCR6+CD3+ T-cells before and after DMF therapy is shown among responders and non-responders from the longitudinal subgroup. Responders exhibit a significant reduction in these cells after therapy, compared to non-responders who do not show a significant change. Interestingly, non-responders start with a slightly higher percentage of CCR6+ T-cells, but this difference does not reach statistical significance in our cohort. (C) The percent change of CCR6+CD3+ T-cells is shown normalized to each patient’s baseline value. It is shown that responders have a significantly higher reduction in peripheral CCR6+CD3+ T-cells compared to non-responders. The lines in the boxplots represent the quartiles of the dataset and the whiskers show the rest of the distribution except for outliers.

### CCR6+ T-cells are the most significant predictors of future disease activity in patients taking dimethylfumarate

Multivariate logistic regression analysis revealed that among various clinical and immunophenotypic metrics from 48 MS patients on long-term treatment with DMF, the percentage of CCR6+CD3+ T-cells was the most significant independent predictor of breakthrough disease activity in the 24 months following the final blood draw (logistic regression controlling for age, sex, race, baseline EDSS, treatment duration, disease duration, FDR-adjusted p = 0.000189) (Table 2). Apart from the CCR6+ percentage of T-cells, other immunophenotypic metrics were significant predictors of disease activity, such as the percentage of CCR6+CD8 T-cells (which includes Tc17 cells), Th17 cells (CD3+CD4+CCR4+CCR6+), Th1-like Th17 cells (CD3+CD4+CCR4-CCR6+) and CCR6+DN T-cells (CD3+CD4-CD8-CCR6+). However, analysis of variance of responders and non-responders revealed that the more specific metrics exhibited increased within group variance and reduced between group variance compared to the CCR6+ proportion of all CD3+ T-cells, suggesting that the more inclusive immunophenotypic metric is more robust. Moreover, the percentage of total T-cells did not significantly predict clinical response, which is in accordance to previous reports^22,23^. The clinical metrics that we used did not reach statistical significance in our logistic regression analysis, except for age and baseline EDSS, thereby indicating a smaller predictive value than immunophenotypic metrics that requires a larger sample size to capture effectively.

### CCR6+ T-cell percentage can effectively discriminate responders to dimethyl-fumarate and stratify the risk of impending disease activity

CCR6+CD3+ T-cell percentage achieved a very high area under the ROC curve (AUC), suggesting that it can effectively discriminate responders and non-responders to DMF (Figure 4A). More specifically, the AUC of CCR6+CD3+ T-cell percentage was 0.85 (95% CI: 0.71–0.99). The AUC of CCR6+CD8+ T-cell percentage, which includes Tc17 cells^5^, was 0.82 (95% CI: 0.69–0.96) and the AUC of Th1.17 cell frequency (CD4+CCR4-CCR6+) was 0.79 (95% CI: 0.64–0.93), suggesting that the more inclusive CCR6+CD3+ percentage could be a superior metric. By using the CCR6+CD3+ threshold obtained by the Youden J statistic^12^ (Youden index = 1.54; threshold = 24.01%; higher percentage was considered positive), we could successfully predict responders to DMF with a very high negative predictive value of 92%. An exploratory analysis of variability in diagnostic accuracy was performed by using a Bayesian bivariate hierarchical model and did not reveal heterogeneity in different patient subgroups of our study. This model estimated a specificity of 0.92 (95% CI: 0.76–0.99) and 0.55 sensitivity (95% CI: 0.23–0.86) for predicting impending disease activity. (Supplementary Figure 2). Finally, based on this cutoff, we performed a cox-proportional hazards analysis, which showed that having high percentage of CCR6+CD3+ T-cells (greater that 24.01%) after at least 3 months of DMF treatment increases the risk of impending disease activity by 11.7 times (hazards ratio 11.7, 95% CI: 2.8–48.3, p = 0.000695), mostly within the upcoming 6 months after the blood draw. (Figure 4B).

**Figure 4.**
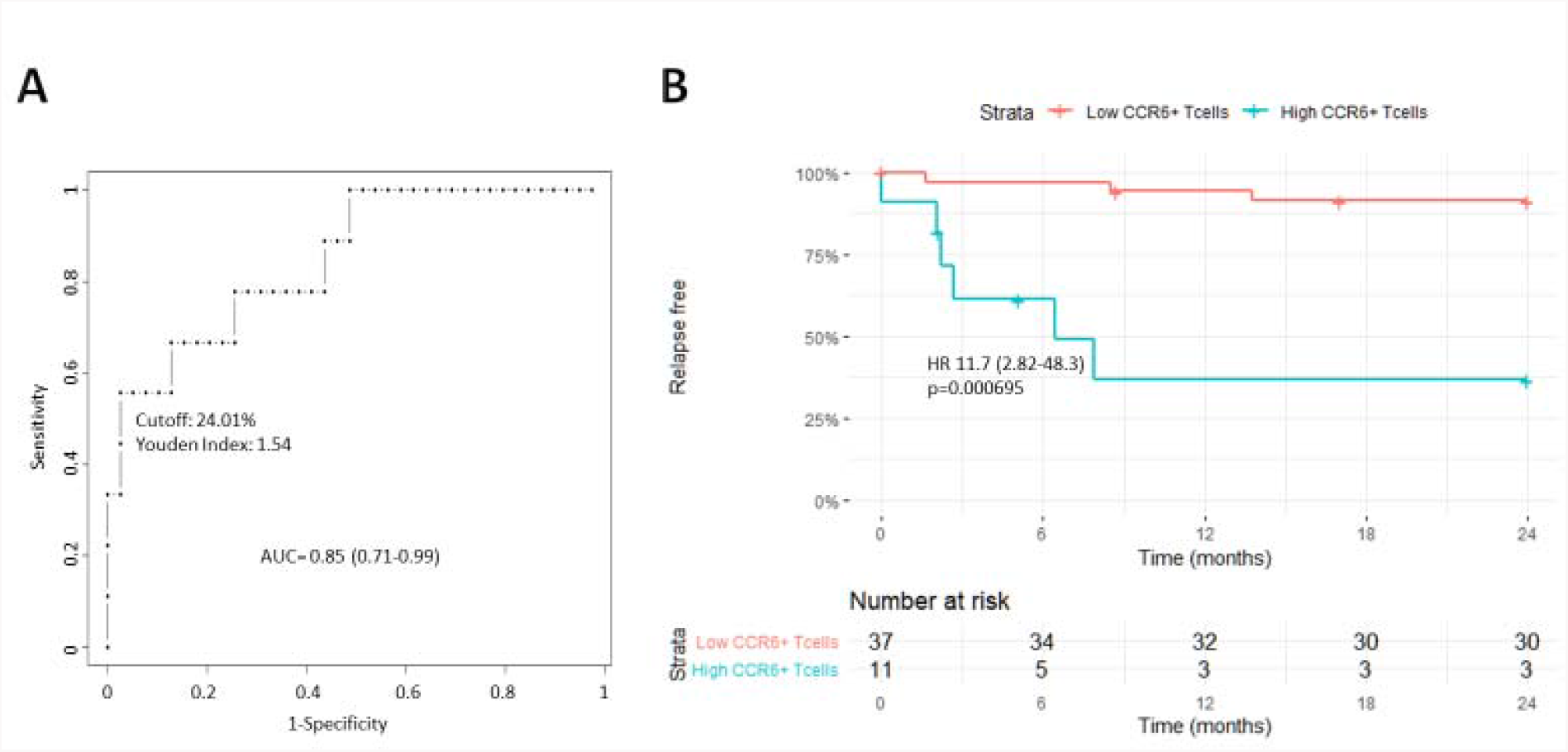
CCR6+ T-cell percentage can discriminate responders and non-responders to DMF therapy and stratify risk of impending disease activity: (A) The discriminative capacity of the percentage of CCR6+CD3+ T-cells was assessed by performing a ROC curve analysis. The area under the curve was 0.85 (95% CI: 0.71–0.99). (B) By using the cutoff threshold identified with the Youden’s J statistic (Youden Index: 1.54; cutoff threshold: 24.01%), patients could be separated into high and low relapse risk groups. Patients with high CCR6+CD3+ T-cell percentage (implying absence of mechanistic response) had 11.7 times higher relapse risk compared to patients with low percentage of those cells (presence of mechanistic response). Patients lost to follow up are marked with a cross on the Kaplan-Meyer curve. Numbers in parenthesis represent 95% confidence intervals.

## Discussion

Currently, initiation of a new MS disease-modifying therapy carries with it a level of uncertainty regarding efficacy at the individual patient level. It is impossible to know whether a patient is responding to a disease-modifying therapy until treatment failure ensues and the patient suffers a clinical relapse, or a new lesion is detected on follow-up MRI. Here we demonstrate the feasibility of predicting treatment response in MS patients by determining the presence or absence of a mechanistic immunophenotypic response from each patient to a specific drug. By measuring the percentage of CCR6+CD3+ T-cells in the peripheral blood of MS patients treated with DMF, we were able to classify patients as responders or non-responders with a high area under the ROC curve. Moreover, by using the cutoff identified by the Youden J statistic, the percentage of CCR6+CD3+ T-cells in each patient could stratify the risk of impending disease activity in the 24 months following their blood draw with a high hazards ratio.

DMF has already been shown to have multiple molecular targets in the peripheral immune system and exert many different biological effects. For example it has been shown to target GADPH and inhibit aerobic glycolysis^24^, activate hydroxyl carboxylic acid receptor 2 (HCAR2)^25^, inhibit NF-κB^26^ and epigenetically modulate microRNAs, such as microRNA-21^1^. One of the effects of DMF we focused on here is the reduction of peripheral CCR6+ T-cells, which has been shown not to be due to increased compartmentalization of CCR6+ T-cells in the CSF^27^. Although CCR6 reduction is not the only immunomodulatory effect of the drug, our findings here reinforce the importance of this effect by DMF, since the CCR6+ percentage of T-cells can separate responders and non-responders to the drug.

Predicting treatment response and risk of future relapse is of great interest to the field of multiple sclerosis and several studies have attempted to address this issue in the past. It is important to make a distinction here between prognostic indicators, which are baseline metrics that can stratify disease severity, and biomarkers of treatment response, which are metrics that can assess response of patients to specific treatments and are the focus of this study^28^. Various methods have been used in the past to predict future disease activity and treatment response^29–36^, from immunophenotyping of peripheral blood^37–40^ and CSF^41^ to cytokine and serologic profiling^42,43^, pharmacogenomics^44^, MRI metrics^45^ and RNA sequencing^38,46^. There have also been several published reports revealing reductions in CCR6+ T-cells in DMF-treated MS patients^1,27,47^, but the relation to treatment response was not evaluated. Regarding treatment response biomarkers for DMF specifically, two studies have used CD8+ T-cell^48^ or IL-17+CD8+ T-cell percentage^49^ and one study used Th1-like Th17 cell frequency^40^. Of note IL17+ CD8 T-cells highly express CCR6^5,50^, which make this population detected in our study (CCR6+CD8+). Based on our data, the more inclusive metric of CCR6+CD3+ T-cell percentage, which includes all of the above cell subtypes in addition to CCR6+DN T-cells (CD3+CD4-CD8-CCR6+) and Th17 cells, provides a more robust biomarker with reduced within group variability and higher AUC. This could be due to the fact that DMF modulates multiple T-cell subtypes in each patient, such as Th17, Th1.17, Tc17 and CCR6+ DN T-cells, thus the CCR6+CD3+ T-cell metric is capturing this more efficiently compared to using only Th1.17 or Tc17 cell frequencies. The inclusion of DN T-cells (CD3+CD4-CD8-CCR6+) in our biomarker is of particular interest since they are produced by chronic T-cell stimulation^51^ and have been shown to remarkably promote neuroinflammation^52^. Finally, the CCR6+CD3+ T-cell percentage is a simple and potentially easily adoptable test, since it only requires a flow cytometer, which is already available in most MS and hematology centers for T and B cell measurements. However, more testing needs to be done to validate the accuracy and precision of this metric across different centers before it is routinely used in clinical practice.

More recently, neurofilament light chain (NFL) has emerged as a validated serologic marker of disease severity and progression^35^. Baseline levels of NFL have significant prognostic utility in MS at the population level^53^, but individual NFL fluctuations do not predict disease activity in the long term^54^, despite the fact that increasing NFL levels can be seen prior to the manifestation of clinical symptoms^55^. Moreover, at the time of increasing NFL levels, damage to the CNS would already be inflicted. By using markers to evaluate peripheral lymphocytes that express brain-homing chemokine receptors in MS patients, such as CCR6+ T-cells, we were able to identify treatment failure with high specificity and stratify the risk of impending disease activity months before it ensued. This suggests that immunologic changes in the periphery of MS patients precede clinical or radiographic disease activity for several months, which is consistent with the peripheral activation model of MS pathogenesis. It is still unclear, whether CCR6 is a marker of treatment response only for DMF or it is a harbinger of disease activity that can be applied to other MS therapeutics. It is also unclear if the addition of other brain-homing cell surface markers, such as CXCR3 and/or α4β7, could potentially improve the predictive power of CCR6^56^. Finally, our results have implications for future personalized medical decision-making strategies in MS. By measuring immunophenotypic changes in each patient before and after treatment initiation, we could separate patients into high and low relapse risk groups early on, after the initiation of therapy and before an MRI is obtained, which can minimize the gap of uncertainty between treatment initiation and determination of clinical response. Based on each patient’s response, a predicted high risk of treatment failure at any point would prompt a change in disease-modifying therapy, potentially avoiding the occurrence of a clinical relapse and/or accumulation of new lesions on imaging. This personalized medical management could potentially minimize unwanted disease activity, accumulation of disability and medication-associated risks, as well as maximize the percentage of patients able to achieve no evidence of disease activity (Figure 5). Clinical trials are needed, however, to test the efficacy of the proposed framework for personalized medical management of MS patients before it can be applied to clinical practice.

**Figure 5.**
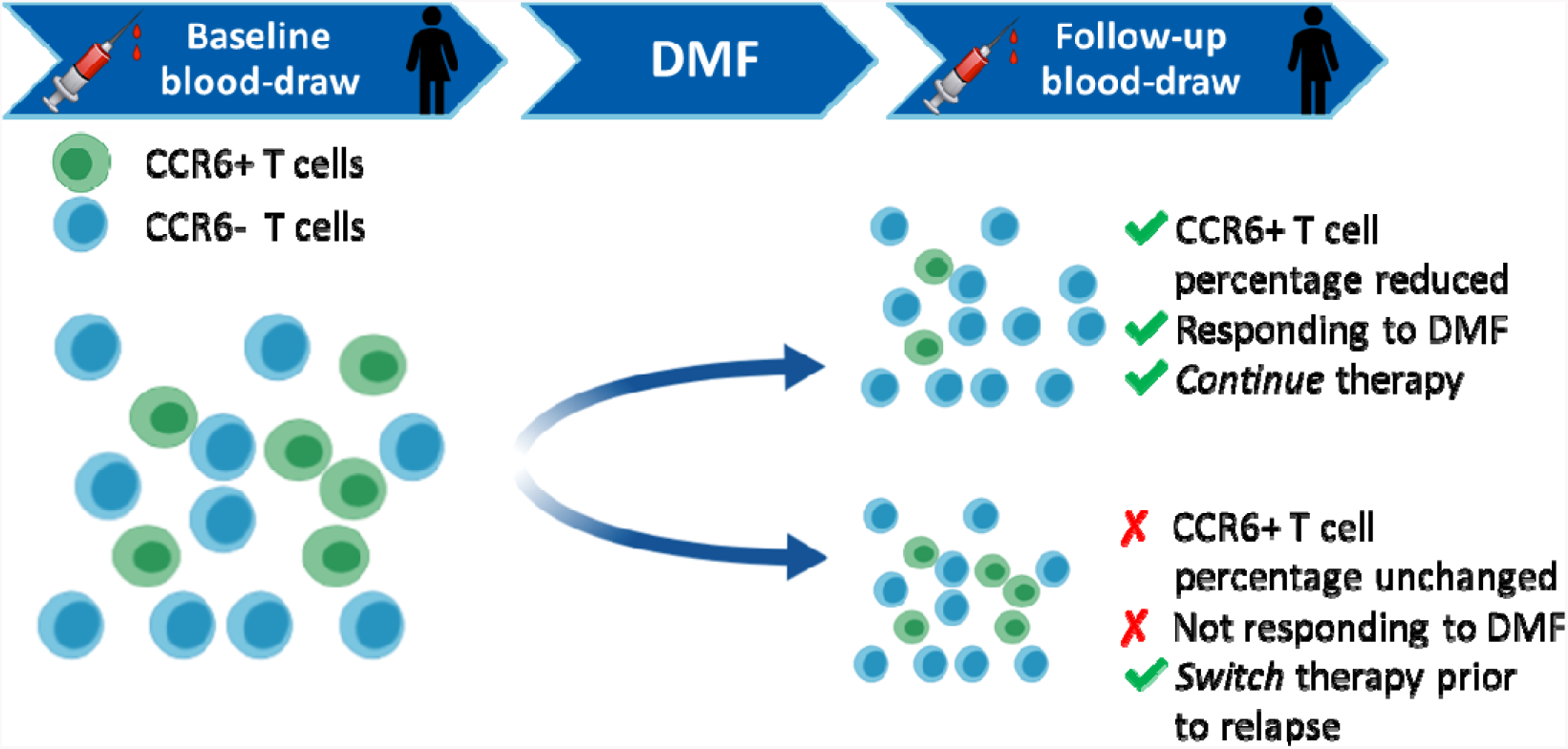
A proposed framework for personalized management in multiple sclerosis: Current treatment escalation strategies in multiple sclerosis are focusing on treatment failure by waiting for breakthrough disease activity to occur either clinically or radiographically on surveillance imaging switching therapy. Here we propose a framework that focuses on identifying treatment success as well as stratifying the risk of treatment failure by utilizing immunophenotypic measurements in peripheral blood of each patient treated with a drug. Absence of immunological changes associated with treatment success s and a predicted high risk of impending disease activity could be used to prompt treatment escalation before the actual development of disease activity. For example, T-cells from different patients will be analyzed for CCR6 positivity at baseline and after treatment with DMF. Lack of reduction of the CCR6+ CD3+ T cell frequency would prompt a change in therapy potentially before a clinical relapse. This biomarker-guided treatment escalation strategy could potentially lead to a higher percentage of no evidence of disease activity (NEDA) among patients with MS and prevent accumulation of disability.

## Data Availability

Data can be provided upon request

## Acknowledgments

The authors would like to thank the human immune core at Mount Sinai, as well as all of those who helped in patient recruitment, especially the MS clinicians and research coordinators at Mount Sinai. Funds were provided by the Friedman Brain Institute and R37-NS42925. There was no involvement of the funding sources to the conduct of this research.

---

**Supplemetal Table 2.** Logistic regression controlling for age, sex, race, baseline EDSS, disease duration and treatment duration. When one of the controlled covariates was the predictor, it was not controlled for in the model. Each row demonstrates the tested predictor for future disease activity in the DMF-treated cohort. FDR-adjusted p-values for different immunophenotypic metrics and clinical characteristics are shown. DN: double negative (CD3+CD4-CD8-), Tc17: T-cytotoxic-17, Th17: T-helper-17, Th1.17: T-helper-1-like T-helper-17.

| Logistic regression | Adjusted P value |
| --- | --- |
| CCR6+CD3+ T cell percentage | **0.000189** |
| CCR6+CD8+ T cell (Tc17) percentage | 0.00754 |
| CCR6+CCR4+CD4+ T cell (Th17) percentage | 0.01685 |
| CCR6+CD4+ T cell percentage | 0.01685 |
| CCR6+CCR4-CD4+ T cell (Th1.17) percentage | 0.02075 |
| CCR6+DN T cell percentage | 0.02075 |
| Age | 0.02518 |
| EDSS | 0.02626 |
| Total CD3- lymphocytes | 0.1588 |
| Total T cell percentage | 0.16637 |
| Treatment Duration | 0.20299 |
| Sex | 0.33197 |
| CCR6+ CD3-CD4-CD8- lymphocyte percentage | 0.5656 |
| Disease Duration | 0.64195 |
| BMI | 0.78477 |
| Race | 0.78477 |

**Supplementary Figure 1.**
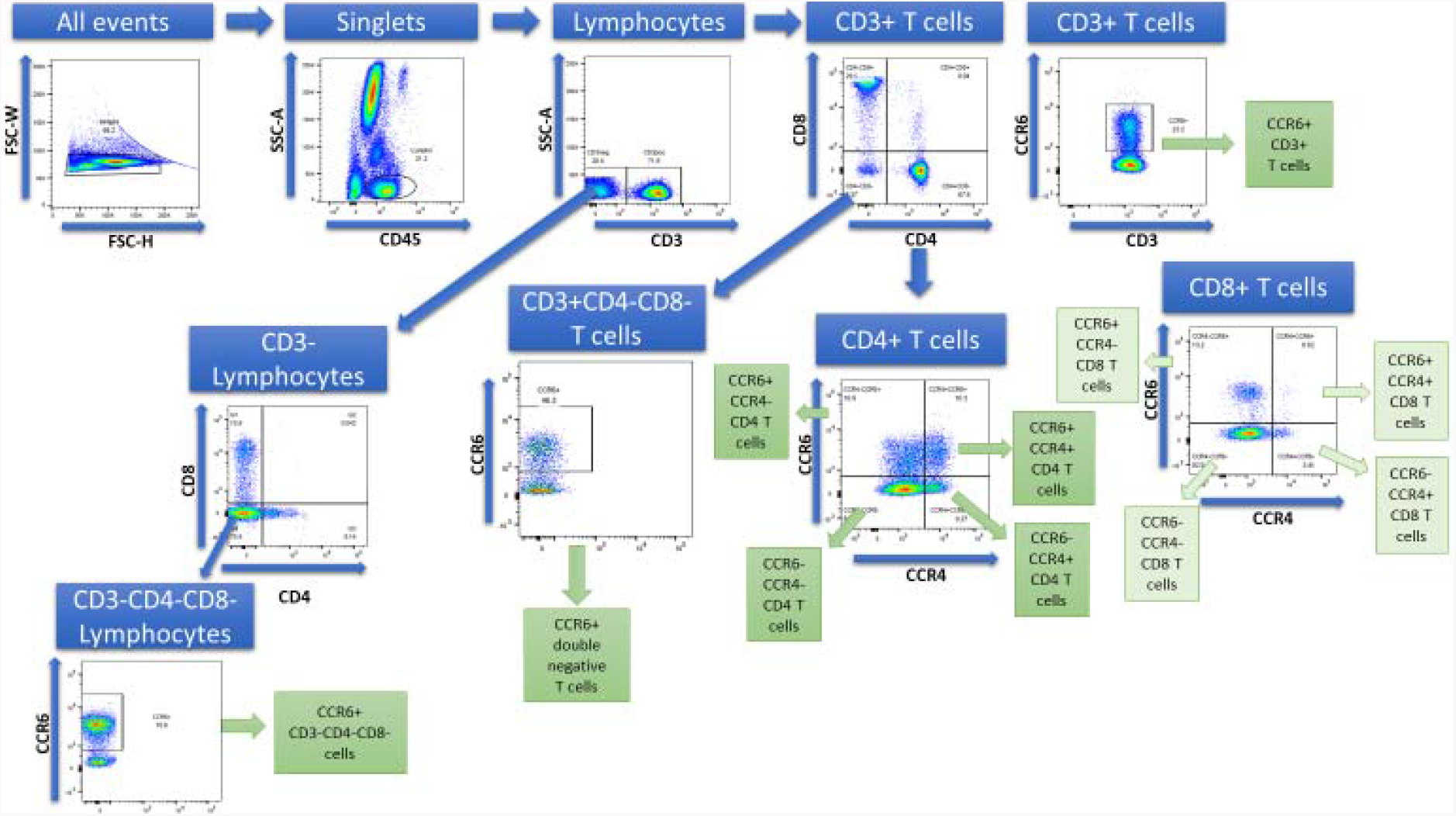
Gating strategy: Doublets were excluded using light scatter measurements and lymphocytes were identified based on their forward and side scatter properties. T-cells were gated using the corresponding cluster of differentiation and/or chemokine receptors and were represented as percentages of the parent population. Initially, all events are gated based on forward scatter (FSC) characteristics to exclude multiplets, thus selecting only singlets (cells in bracket in first figure). The singlets are further gated by complexity/granularity of cells based on side scatter characteristics (SSC). Lymphocytes are circled in the second image and are discriminated from monocytes (medium SSC values) and granulocytes (high SSC values) based on their very low SSC values. T-cells are gated based on CD3 positivity. The CD3 negative population is further gated to only include CD4 negative and CD8 negative cells and thus mostly include B cells. CCR6 positive percentage of this CD3-CD4-CD8-population is then obtained. CD3+ T-cells are further gated using CD4 and CD8 markers and helper(CD4+), cytotoxic (CD8+) and double negative (CD4-CD8-) T-cells are obtained. CCR6 and CCR4 positive percentages of these T-cells are obtained in the next gates.

**Supplementary Figure 2.**
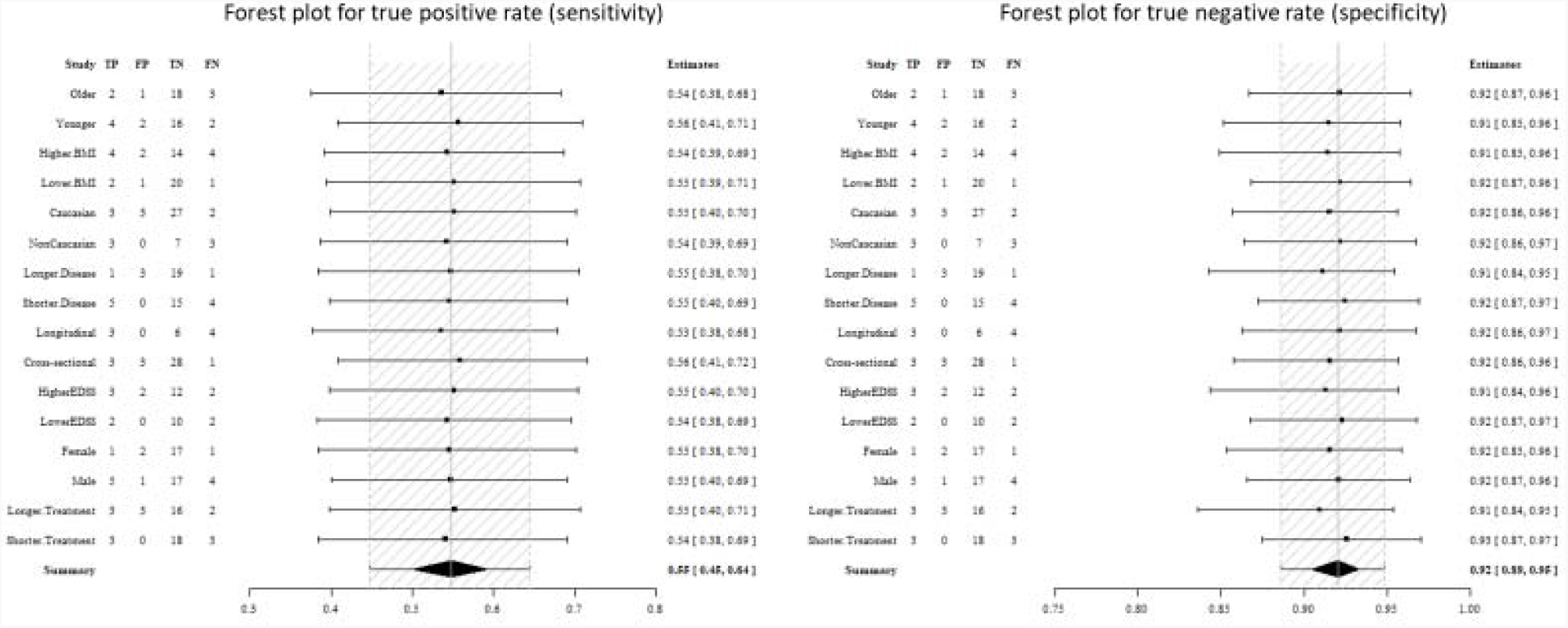
Exploratory analysis of variability in diagnostic accuracy: Patients were separated into subgroups based on their demographic characteristics. Continuous variables were transformed into categorical variables based on having a value higher or lower than the median. A Bayesian bivariate hierarchical model was used to estimate sensitivity and specificity of the test in these subgroups and revealed similar results for all of them.

## References

1 Ntranos A, Ntranos V, Bonnefil V, et al. Fumarates target the metabolic-epigenetic interplay of brain-homing T cells in multiple sclerosis. Brain 2019; 142: 647–61.

2 Reboldi A, Coisne C, Baumjohann D, et al. C-C chemokine receptor 6-regulated entry of TH-17 cells into the CNS through the choroid plexus is required for the initiation of EAE. Nat Immunol 2009; 10: 514–23.

3 Liston A, Kohler RE, Townley S, et al. Inhibition of CCR6 Function Reduces the Severity of Experimental Autoimmune Encephalomyelitis via Effects on the Priming Phase of the Immune Response. J Immunol 2009; 182: 3121–30.

4 Wang C, Kang SG, Lee J, Sun Z, Kim CH. The roles of CCR6 in migration of Th17 cells and regulation of effector T-cell balance in the gut. Mucosal Immunol 2009; 2: 173–83.

5 Saxena A, Desbois S, Carrié, N, Lawand M, Mars LT, Liblau RS. Tc17 CD8 + T Cells Potentiate Th1-Mediated Autoimmune Diabetes in a Mouse Model. J Immunol 2012; 189: 3140–9.

6 Cao Y, Goods BA, Raddassi K, et al. Functional inflammatory profiles distinguish myelin-reactive T cells from patients with multiple sclerosis. Sci Transl Med 2015; 7: 287ra74-287ra74.

7 Ramesh R, Kozhaya L, McKevitt, K, et al. Pro-inflammatory human Th17 cells selectively express P-glycoprotein and are refractory to glucocorticoids. J Exp Med 2014; 211: 89–104.

8 Huber M, Heink S, Pagenstecher A, et al. IL-17A secretion by CD8+T cells supports Th17-mediated autoimmune encephalomyelitis. J Clin Invest 2013; 123: 247–60.

9 Polman CH, Reingold SC, Banwell B, et al. Diagnostic criteria for multiple sclerosis: 2010 Revisions to the McDonald criteria. Ann Neurol 2011; 69: 292–302.

10 Giovannoni G, Tomic D, Bright JR, Havrdová, E. “No evident disease activity”: The use of CCR6+ T-cells as a biomarker of DMF response in MS combined assessments in the management of patients with multiple sclerosis. Mult. Scler. 2017; 23: 1179–87.

11 Dobson R, Rudick RA, Turner B, Schmierer K, Giovannoni G. Assessing treatment response to interferon-:Is there a role for MRI? Neurology 2014; 82: 248–54.

12 Youden WJ. Index for rating diagnostic tests. Cancer 1950; 3: 32–5.

13 Thomas L. Retrospective power analysis. Conserv Biol 1997; 11: 276–80.

14 Fairweather PG. Statistical power and design requirements for environmental monitoring. Mar Freshw Res 1991; 42: 555–67.

15 Obuchowski NA, Lieber ML, Wians FH. ROC curves in Clinical Chemistry: Uses, misuses, and possible solutions. Clin. Chem. 2004; 50: 1118–25.

16 Robin X, Turck N, Hainard A, et al. pROC: an open-source package for R and S+ to analyze and compare ROC curves. BMC Bioinformatics 2011; 12: 77.

17 Benjamini Y, Hochberg Y. Controlling the False Discovery Rate: A Practical and Powerful Approach to Multiple Testing. J R Stat Soc Ser B 1995; 57: 289–300.

18 Therneau, Terry M., Grambsch PM. Modeling Survival Data: Extending the Cox Model. Technometrics 2009; 44: 85–6.

19 Campbell JJ, O’Connell, DJ, Wurbel M-A. Cutting Edge: Chemokine Receptor CCR4 Is Necessary for Antigen-Driven Cutaneous Accumulation of CD4 T Cells under Physiological Conditions. J Immunol 2007; 178: 3358–62.

20 Gold R, Kappos L, Arnold DL, et al. Placebo-Controlled Phase 3 Study of Oral BG-12 for Relapsing Multiple Sclerosis. N Engl J Med 2012; 367: 1098–107.

21 Fox RJ, Miller DH, Phillips JT, et al. Placebo-controlled phase 3 study of oral BG-12 or glatiramer in multiple sclerosis. N Engl J Med 2012; 367: 1087–97.

22 Fox RJ, Chan A, Gold R, et al. Characterizing absolute lymphocyte count profiles in dimethyl fumarate’treated patients with MS Patient management considerations. Neurol Clin Pract 2016; 6: 220–9.

23 Ghadiri M, Rezk A, Li R, et al. Dimethyl fumarate-induced lymphopenia in MS due to differential T-cell subset apoptosis. Neurol Neuroimmunol NeuroInflammation 2017; 4: e340.

24 Sui P, Wiesner DL, Xu J, et al. Dimethyl fumarate targets GAPDH and aerobic glycolysis to modulate immunity. Science (80-.). 2018;: 1–16.

25 Chen H, Assmann JC, Krenz A, et al. Hydroxycarboxylic acid receptor 2 mediates dimethyl fumarate’s protective effect in EAE. J Clin Invest 2014; 124: 2188–92.

26 Gillard GO, Collette B, Anderson J, et al. DMF, but not other fumarates, inhibits NF-κB activity in vitro in an Nrf2-independent manner. J Neuroimmunol 2015; 283: 74–85.

27 Holm Hansen R, Højsgaard Chow H, Christensen JR, Sellebjerg F, von Essen MR. Dimethyl fumarate therapy reduces memory T cells and the CNS migration potential in patients with multiple sclerosis. Mult Scler Relat Disord 2020; 37. DOI:10.1016/j.msard.2019.101451.

28 Gasperini C, Prosperini L, Tintoré, M, et al. Unraveling treatment response in multiple sclerosis: A clinical and MRI challenge. Neurology. 2019; 92: 180–92.

29 Quirant-Sánchez, B, Hervás-García, J V., Teniente-Serra, A, et al. Predicting therapeutic response to fingolimod treatment in multiple sclerosis patients. CNS Neurosci Ther 2018; 24: 1175–84.

30 Lorefice L, Murgia F, Fenu G, et al. Assessing the Metabolomic Profile of Multiple Sclerosis Patients Treated with Interferon Beta 1a by 1H-NMR Spectroscopy. Neurotherapeutics 2019;: 1–11.

31 Paolicelli D, Manni A, D’Onghia, M, et al. Lymphocyte subsets as biomarkers of therapeutic response in Fingolimod treated Relapsing Multiple Sclerosis patients. J Neuroimmunol 2017; 303: 75–80.

32 Río, J, Rovira, À, Tintoré, M, et al. Disability progression markers over 6–12 years in interferon-β treated multiple sclerosis patients. Mult Scler J 2018; 24: 322–30.

33 Río, J, Auger C, Rovira, À. MR Imaging in Monitoring and Predicting Treatment Response in Multiple Sclerosis. Neuroimaging Clin. N. Am. 2017; 27: 277–87.

34 Medina S, Villarrubia N, Sainz de la Maza S, et al. Optimal response to dimethyl fumarate associates in MS with a shift from an inflammatory to a tolerogenic blood cell profile. Mult Scler J 2018; 24: 1317–27.

35 Kuhle J, Kropshofer H, Haering DA, et al. Blood neurofilament light chain as a biomarker of MS disease activity and treatment response. Neurology 2019; 92: e1007–15.

36 Valenzuela RM, Kaufman M, Balashov KE, Ito K, Buyske S, Dhib-Jalbut, S. Predictive cytokine biomarkers of clinical response to glatiramer acetate therapy in multiple sclerosis. J Neuroimmunol 2016; 300: 59–65.

37 Posová, H, Horáková, D, Čapek, V, Uher T, Hrušková, Z, Hrušková, Z Havrdová, E. Peripheral blood lymphocytes immunophenotyping predicts disease activity in clinically isolated syndrome patients. BMC Neurol 2017 17: 145.

38 Moreno-Torres, I, González-García, C, Marconi M, et al. Immunophenotype and transcriptome profile of patients with multiple sclerosis treated with fingolimod: Setting up a model for prediction of response in a 2-year translational study. Front Immunol 2018; 9: 1693.

39 Alvarez E, Parks BJ, Piccio L, et al. Predicting optimal response to B-cell depletion with rituximab in multiple sclerosis using CXCL13 index, magnetic resonance imaging and clinical measures. Mult Scler J – Exp Transl Clin 2015; 1: 205521731562380.

40 Mansilla MJ, Navarro-Barriuso J, Presas-Rodríguez S, et al. Optimal response to dimethyl fumarate is mediated by a reduction of Th1-like Th17 cells after 3 months of treatment. CNS Neurosci Ther 2019; 25: 995–1005.

41 Ohayon J, Toro C, Romm E, et al. Comprehensive Immunophenotyping of Cerebrospinal Fluid Cells in Patients with Neuroimmunological Diseases. J Immunol 2014; 192: 2551–63.

42 Moţăţăţianu, A, Andone S, Brcuean, LI, et al. Clinical and Serological Biomarkers of Treatment’s Response in Multiple Sclerosis Patients Treated Continuously with Interferonβ-1b for More than a Decade. CNS Neurol Disord – Drug Targets 2018; 17: 780–92.

43 Buck D, Hemmer B. Biomarkers of treatment response in multiple sclerosis. Expert Rev. Neurother. 2014; 14: 165–72.

44 Coyle PK. Pharmacogenetic Biomarkers to Predict Treatment Response in Multiple Sclerosis: Current and Future Perspectives. Mult Scler Int 2017; 2017: 1–10.

45 Tur C, Nos C, Otero-Romero, S, et al. Disability progression markers over 6–12 years in interferon-β-treated multiple sclerosis patients. Mult Scler J 2017; 24: 322–30.

46 Hecker M. Blood transcriptome profiling captures dysregulated pathways and response to treatment in neuroimmunological disease. EBioMedicine. 2019; 49: 2–3.

47 Mehta D, Miller C, Arnold DL, et al. Effect of dimethyl fumarate on lymphocytes in RRMS: Implications for clinical practice. Neurology 2019; 92: e1724–38.

48 Fleischer V, Friedrich M, Rezk A, et al. Treatment response to dimethyl fumarate is characterized by disproportionate CD8+ T cell reduction in MS. Mult Scler J 2018; 24: 632–41.

49 Lückel, C, Picard F, Raifer H, et al. IL-17+ CD8+ T cell suppression by dimethyl fumarate associates with clinical response in multiple sclerosis. Nat Commun 2019; 10: 5722.

50 Oo YH, Banz V, Kavanagh D, et al. CXCR3-dependent recruitment and CCR6-mediated positioning of Th-17 cells in the inflamed liver. J Hepatol 2012; 57: 1044–51.

51 Grishkan I V., Ntranos A, Calabresi PA, Gocke AR. Helper T cells down-regulate CD4 expression upon chronic stimulation giving rise to double-negative T cells. Cell Immunol 2013; 284: 68–74.

52 Meng H, Zhao H, Cao X, et al. Double-negative T cells remarkably promote neuroinflammation after ischemic stroke. Proc Natl Acad Sci U S A 2019; 116: 5558–63.

53 Berger T, Stüve, O. Neurofilament light chain. Neurology 2019; 92: 10.1212/WNL.0000000000007022.

54 Cantó, E, Barro C, Zhao C, et al. Association between Serum Neurofilament Light Chain Levels and Long-term Disease Course among Patients with Multiple Sclerosis Followed up for 12 Years. JAMA Neurol 2019; 76: 1359–66.

55 Akgün K, Kretschmann N, Haase R, et al. Profiling individual clinical responses by high-frequency serum neurofilament assessment in MS. Neurol Neuroimmunol NeuroInflammation 2019; 6: e555.

56 Van Langelaar J, Van Der Vuurst De Vries RM, Janssen M, et al. T helper 17.1 cells associate with multiple sclerosis disease activity: Perspectives for early intervention. Brain. 2018; 141: 1334–49.

